# Understanding Acute Respiratory Illness in Nepalese Children Under Five: Insights from the 2016 NDHS

**DOI:** 10.1101/2025.02.11.25322058

**Authors:** Anish Dhodari, Anisha Dhakal, Joshna Pokhrel, Pragya Bhattarai

## Abstract

Acute Respiratory Illness (ARI) remains a critical public health challenge for children under five in Nepal, with persistent socioeconomic inequities in disease burden. This cross-sectional study analyzed Nepal’s 2016 Demographic and Health Survey (n=4,887) to identify determinants of ARI using caregiver reported symptoms of rapid breathing and chest-related issues within two weeks of the survey. Weighted logistic regression analyses examined associations between ARI and variables such as wealth quintile, residence type, maternal education, and child age.

Nationally, ARI prevalence stood at 2.41%, with pronounced disparities: children from the poorest wealth quintile had 5.6× higher prevalence than the richest (3.32% vs 0.59%), while Province 6 reported the highest regional burden (3.37%). Multivariate analysis confirmed household wealth as the significant predictor (AOR=1.60, 95% CI:0.063–40.836), though wide confidence intervals indicated model overdispersion. Notably, 84.92% of caregivers sought healthcare—primarily through private facilities (32.30%) and pharmacies (27.92%)—highlighting gaps in public sector utilization

These findings reveal wealth-based health disparities despite Nepal’s progress toward SDG child mortality targets. The results advocate for province-specific interventions in high-burden regions like Province 6, including mobile pneumonia clinics and biomass fuel transition programs. Socioeconomic disparities play a key role in ARI prevalence among children under five in Nepal. Interventions aimed at reducing care-seeking fragmentation while targeting poverty-alleviation strategies to reduce ARI’s disproportionate impact on marginalized populations could be beneficial. Future research should focus on identifying additional risk factors that were not looked into by the NDHS survey and evaluating targeted prevention strategies.

## Introduction

Acute respiratory illness (ARI) remains a significant global health challenge, particularly for children under five years of age.^1 2^ Defined by the World Health Organization as respiratory illnesses causing cough, fever, and hospitalization within 10 days of onset ^3^, ARI accounts for approximately 1.3 million deaths annually in children under five, with two-thirds occurring in infants.^4^ Additionally, ARI leads to over 12 million hospital admissions globally among this age group each year.^5^ This burden is particularly pronounced in developing countries, where limited access to healthcare and resources exacerbates the issue. ^6 7^

In South Asia, ARI contributes to 15% of childhood mortality.^8^ Bangladesh Demographic and Health Survey data revealed a 35.8% prevalence of ARI among children under five, with risk factors including age, delivery method, family size, education, and regional disparities.^9^ Nepal faces similar challenges, with ARI constituting a substantial proportion of hospital visits and admissions for children under five.^10 11^ A study in Kathmandu Valley reported ARI symptoms in 60.8% of children sampled from tertiary hospitals.^10^ Environmental factors, such as air pollution, overcrowding, and poor sanitation, significantly exacerbate ARI risk, especially in resource-constrained settings similar to Nepal and many South Asian countries. ^12 13^ Despite this, Nepal has achieved relatively low under-five mortality rates—28 deaths per 1,000 live births^14^ —and aims to reduce this to 25 by 2030 in line with the Sustainable Development Goals.^15^

Key contributors to ARI include viral pathogens such as respiratory syncytial virus (RSV) and influenza virus, as well as bacterial agents like *Streptococcus pneumoniae* and *Haemophilus influenzae* type b.^16^ Additionally, household reliance on solid fuels like kerosene and wood for cooking further increases the vulnerability of young children to respiratory infections. Socio-economic determinants, including household wealth, maternal education, and access to healthcare, play a critical role in influencing ARI prevalence and outcomes.^17^

This study leverages data from the 2016 Nepal Demographic and Health Survey (NDHS) to examine the prevalence of ARI among children under five. This study fills an evidence gap by analyzing how maternal education, household wealth, and residence type influence ARI prevalence and healthcare-seeking behavior in Nepal.. The findings will not only provide valuable insights into the specific drivers of ARI in Nepal but also contribute to broader global health efforts aimed at reducing child mortality and morbidity from respiratory illnesses. Ultimately, this study aims to inform public health interventions and policies to address ARI, contributing to the global agenda of improving child health and achieving the Sustainable Development Goals (SDGs) for child health.

## Materials AND Methods

### Data Source

This study utilized data from the 2016 Nepal Demographic and Health Survey (NDHS), a nationally representative survey conducted by the Ministry of Health and Population, Nepal, was implemented by New ERA. The NDHS employed a two-stage stratified cluster sampling design to ensure comprehensive coverage of Nepal’s urban and rural populations across all seven provinces. Detailed survey methodologies, including sampling procedures, are outlined in the NDHS 2016 report.^18^ The data for the NDHS 2016 was obtained in March of 2023. The Authors had no access to information that could potentially identify individual participants from the study.

### Study Population

The target population for this study included children under five years of age. ARI status was determined based on caregiver-reported symptoms of rapid breathing and chest-related issues in the two weeks preceding the survey. After excluding cases with incomplete information on ARI symptoms or key covariates, the final analytic sample comprised 4,887 children. Survey weights were applied to account for the complex sampling design and ensure nationally representative estimates.

### Variables and Measurements

The primary outcome variable for this study was **Acute Respiratory Illness (ARI)**. ARI status was coded as a binary variable: “1” for children with symptoms of ARI and “0” for those without.

### Explanatory variables

were selected based on existing literature and relevance to the study objectives. These included:

1. **Child’s Sex**: A binary variable categorized as male or female.
2. **Child’s Age**: A categorical variable created by grouping children into five age categories: 0–11 months, 12–23 months, 24–35 months, 36–47 months, and 48–59 months.
3. **Household Wealth Quintile**: An ordinal variable representing household socioeconomic status, categorized as poorest, poorer, middle, richer, and richest, based on asset ownership, housing materials, and access to utilities.
4. **Maternal Education Level**: An ordinal variable categorized as no education, primary education, secondary education, or higher education.
5. **Type of Residence**: A binary variable classified as urban or rural.
6. **Province**: A nominal categorical variable representing the administrative divisions of Nepal, with seven categories corresponding to the seven provinces: Province 1, Province 2, Province 3, Province 4, Province 5, Province 6, and Province 7.

All variables were cleaned, recorded, and labeled as necessary to ensure consistency and clarity.

### Statistical Analysis

The analysis involved descriptive, bivariate, and multivariate statistical approaches to address the study objectives. Weighted descriptive statistics were used to calculate the prevalence of Acute Respiratory Illness (ARI) and to summarize the socio-demographic characteristics of children under five years of age. Bivariate associations between ARI and explanatory variables, including child sex, age, maternal education, household wealth, type of residence, and province, were assessed using chi-squared tests. Variables with a p-value < 0.05 were considered statistically significant and included in the multivariable analysis. Multivariate logistic regression models were used to identify independent predictors of ARI. The models were adjusted for potential confounders to estimate adjusted odds ratios (AORs) and their corresponding 95% confidence intervals (CIs). Both unadjusted and adjusted models were presented to highlight crude associations and the independent effects of explanatory variables. All analyses were performed using Stata version 17.

### Ethics Statement

This study is based on publicly available, de-identified secondary data from the 2016 Nepal Demographic and Health Survey (NDHS). The NDHS was conducted by the Nepal Ministry of Health and Population in collaboration with ICF International and New ERA, with ethical approval obtained from the Nepal Health Research Council (NHRC). Informed consent was obtained from all survey participants by NDHS before data collection. As this study involved secondary data analysis with no direct interaction with human participants, additional ethical approval was not required.

## Results

The weighted prevalence of Acute Respiratory Illness (ARI) among children under five years of age in Nepal was 2.41% (118 out of 4,887 children). Males comprised 52.45% of the sample, and children aged 12–23 months represented the largest age group (21.45%). Nearly half (45.80%) resided in rural areas, and 34.04% of mothers had no formal education. Household wealth distribution revealed that 21.29% of children belonged to the poorest quintile, while 14.99% were from the richest quintile. Among children with ARI symptoms, 84.92% were taken to a healthcare provider for advice or treatment. Private medical facilities (32.30%) and pharmacies (27.92%) were the most utilized healthcare providers, while 15.08% of caregivers did not seek care (Table 2).

**Table 1:** Prevalence of Acute Respiratory Illness (ARI) Among Children Under Five in Nepal.

| ARI Status | Weighted Frequency | Weighted Percentage |
| --- | --- | --- |
| No ARI | 4769 | 97.59 |
| ARI | 118 | **2.41** |
| Total | 4887 | 100.00 |
| **Prevalence = 2.41%** |  |  |

**Table 2:**
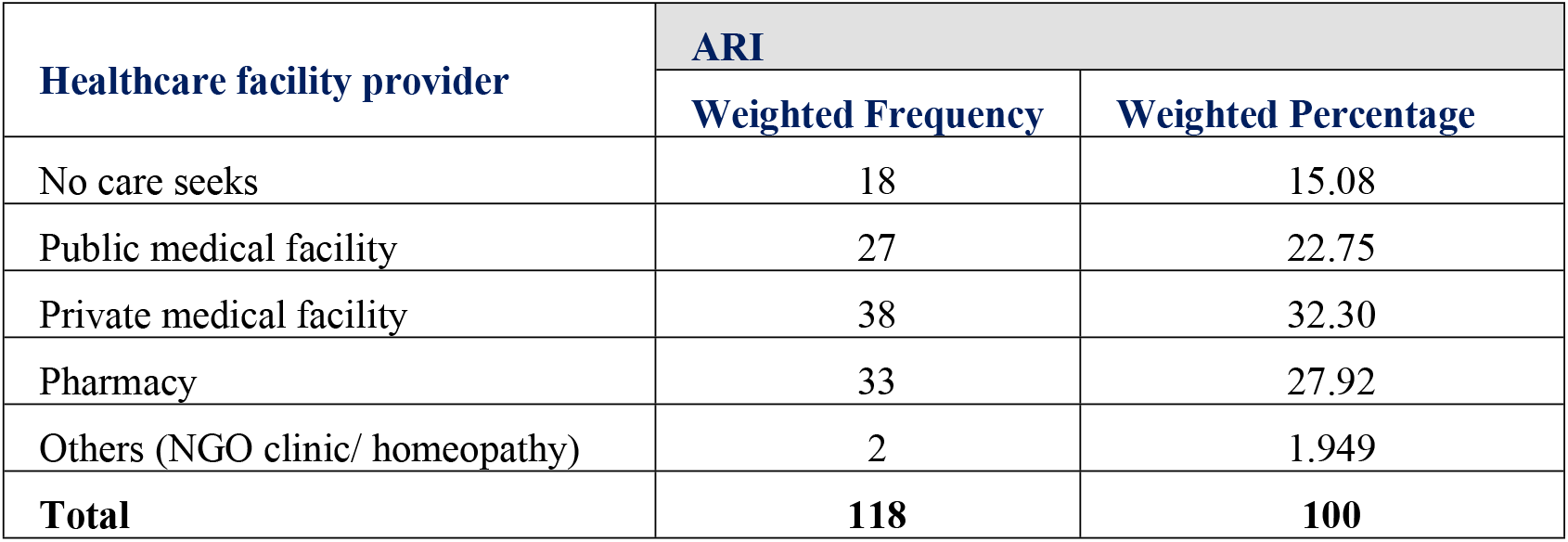
Healthcare advice sought for children under 5 years of age with ARI.

| Healthcare facility provider | ARI |  |
| --- | --- | --- |
|  | Weighted Frequency | Weighted Percentage |
| No care seeks | 18 | 15.08 |
| Public medical facility | 27 | 22.75 |
| Private medical facility | 38 | 32.30 |
| Pharmacy | 33 | 27.92 |
| Others (NGO clinic/ homeopathy) | 2 | 1.949 |
| **Total** | **118** | **100** |

Bivariate analysis (Table 3) identified **household wealth quintile** as the only variable significantly associated with ARI (p = 0.008). Children from the poorest households had the highest ARI prevalence (3.32%), compared to 0.59% among the richest. No significant associations were observed for child sex, age, maternal education, residence type, or province. Multivariate logistic regression (Table 4) confirmed the persistence of wealth-based disparities. Children from the poorest households had **1.60 times higher odds** of ARI compared to the richest (AOR: 1.60; 95% CI: 0.063–40.836). However, this association lacked precision, as reflected by the wide confidence interval. Other variables, including province and maternal education, showed no significant association with ARI in the adjusted model.

**Table 3:** Bivariate Associations Between Socio-Demographic Factors and ARI Among Children Under Five in Nepal.

| Factors | No ARI (%) | ARI (%) | Chi-squared Value | P-value |
| --- | --- | --- | --- | --- |
| Sex of Child |  |  |  |  |
| Male | 97.34 | 2.66 | 1.45 | 0.293 |
| Female | 97.87 | 2.13 |  |  |
| Age of Child (in months) |  |  |  |  |
| 0-11 | 96.35 | 3.65 | 10.295 | 0.602 |
| 12-23 | 96.58 | 3.42 |  |  |
| 24-35 | 96.7 | 3.3 |  |  |
| 36-47 | 98.74 | 1.26 |  |  |
| 48-59 | 98.58 | 1.42 |  |  |
| Type of Residence |  |  |  |  |
| Urban | 97.92 | 2.077 | 2.738 | 0.146 |
| Rural | 97.19 | 2.807 |  |  |
| Province |  |  |  |  |
| Province1 | 96.69 | 3.31 | 10.044 | 0.183 |
| Province2 | 98.49 | 1.51 |  |  |
| Province3 | 97.64 | 2.36 |  |  |
| Province4 | 98.28 | 1.73 |  |  |
| Province5 | 97.15 | 2.85 |  |  |
| Province6 | 96.63 | 3.37 |  |  |
| Province7 | 97.4 | 2.6 |  |  |
| Mother’s Education Level |  |  |  |  |
| No education | 97.88 | 2.12 | 2.172 | 0.604 |
| Primary | 97.18 | 2.82 |  |  |
| Secondary | 97.35 | 2.65 |  |  |
| Higher Secondary | 98.02 | 1.98 |  |  |
| Wealth Index |  |  |  |  |
| Poorest | 96.69 | 3.32 | 16.617 | 0.008 |
| Second | 96.88 | 3.12 |  |  |
| Middle | 97.57 | 2.43 |  |  |
| Fourth | 97.95 | 2.05 |  |  |
| Richest | 99.41 | 0.59 |  |  |
| Currently Breastfeeding |  |  |  |  |
| Yes | 97.49 | 2.51 | 0.614 | 0.451 |
| No | 97.9 | 2.1 |  |  |
| Status of Employment |  |  |  |  |
| No | 49.66 | 1 | 3.901 | 0.088 |
| ARI | 47.93 | 1.41 |  |  |
| Vaccination Status |  |  |  |  |
| Yes | 97.67 | 2.33 | 0.141 | 0.689 |
| No | 97.14 | 2.86 |  |  |

**Table 4:** Multivariate Logistic Regression Analysis of Factors Associated with ARI Among Children Under Five in Nepal.

| Variables | Odds ratio (OR) | P-value | 95% CI of OR |  |
| --- | --- | --- | --- | --- |
|  |  |  | Lower | Upper |
| Sex of Child |  |  |  |  |
| Female | Reference |  |  |  |
| Male | 1.06 | 0.91 | 0.413 | 2.697 |
| Age of child(months) |  |  |  |  |
| 36-37 | Reference |  |  |  |
| 0-11 | 0.67 | 0.701 | 0.085 | 5.251 |
| 12-23 | 1.75 | 0.269 | 0.647 | 4.740 |
| 24-35 | 1 | (omitted) |  |  |
| 48-59 | 1 | (empty) |  |  |
| Province |  |  |  |  |
| Province 1 | Reference |  |  |  |
| Province 2 | 0.91 | 0.928 | 0.113 | 7.321 |
| Province 3 | 7.54 | 0.066 | 0.874 | 65.035 |
| Province 4 | 4.03 | 0.259 | 0.356 | 45.597 |
| Province 5 | 4.16 | 0.131 | 0.651 | 26.654 |
| Province 6 | 12.72 | 0.013 | 1.725 | 93.721 |
| Province 7 | 5.16 | 0.117 | 0.659 | 40.441 |
| Residence |  |  |  |  |
| Urban | Reference |  |  |  |
| Rural | 0.72 | 0.416 | 0.323 | 1.597 |
| Wealth index |  |  |  |  |
| Richest | Reference |  |  |  |
| Poorest | 1.60 | 0.774 | 0.063 | 40.836 |
| Poorer | 3.64 | 0.405 | 0.172 | 77.193 |
| Middle | 4.82 | 0.322 | 0.213 | 108.758 |
| Richer | 2.57 | 0.557 | 0.109 | 60.361 |
| **Education** |  |  |  |  |
| No education | *Reference* |  |  |  |
| Primary | 0.99 | 0.993 | 0.263 | 3.756 |
| Secondary | 0.41 | 0.167 | 0.118 | 1.450 |
| Higher | 0.39 | 0.351 | 0.054 | 2.827 |
| **Breast feeding** |  |  |  |  |
| No | *Reference* |  |  |  |
| Yes | 1 | Empty |  |  |
| **Working Status** |  |  |  |  |
| No | *Reference* |  |  |  |
| Yes | 1.32 | 0.601 | 0.462 | 3.793 |
| **Vaccination status** |  |  |  |  |
| Not vaccinated | *Reference* |  |  |  |
| Vaccinated | 0.33 | 0.251 | 0.050 | 2.199 |

Provincial differences in ARI prevalence were notable but not statistically significant in either the bivariate or multivariate analyses. The highest prevalence occurred in **Province 6 (3.37%)** and **Province 1 (3.31%)**, while the lowest was in Province 2 (1.51%).

## Discussion

The prevalence of ARI was 2.41%, indicating improvements in child health compared to SouthAsian counterparts but highlighting the continued vulnerability of certain subpopulations to preventable respiratory illnesses. However, the 2022 NDHS reported a further decline in ARI prevalence to 1%, suggesting continued progress in reducing the burden of respiratory infections among children under five.^19^

Healthcare-seeking behavior among caregivers was encouraging in this study, with 84.92% of children with ARI symptoms receiving medical advice or treatment (Table 2). However, the 2022 NDHS reported a slightly lower rate of care-seeking at 75%, indicating a potential decline in healthcare utilization for ARI cases.^19^ This study found that private medical facilities (32.30%) and pharmacies (27.92%) were the most utilized healthcare providers for ARI treatment. The NDHS 2022 reported an even greater reliance on private sector healthcare, with 60% of ARI cases managed in private clinics and pharmacies. A study done in Bangladesh showed that most caregivers sought medical care for children with ARI symptoms, but the reliance on private facilities may lead to financial challenges and concerns about care quality.^20^ Public healthcare systems must be strengthened to provide accessible, affordable, and standardized care for ARI to reduce inequities in health outcomes.

A significant association was observed between household wealth quintile and ARI prevalence (Table 3). Children from the poorest households had the highest prevalence (3.32%), while those from the richest households had the lowest prevalence (0.59%). Global evidence supports the link between poverty and increased respiratory illness risk, driven by factors such as reliance on biomass fuels, poor housing, and limited access to healthcare.^21^ Interventions like promoting clean cooking technologies, improving housing conditions, and poverty alleviation programs are essential to mitigating ARI risk among socioeconomically disadvantaged populations.

Even after adjusting for confounders, children from poorer households exhibited higher odds of ARI compared to their wealthier counterparts (AOR: 1.60; 95% CI: 0.063–40.836) (Table 4). These findings align with global and regional evidence linking poverty to increased susceptibility to respiratory illnesses due to factors such as reliance on biomass fuels, poor housing conditions, and limited access to healthcare services. Targeted interventions, such as clean cooking technologies, housing improvements, and poverty alleviation programs, are essential to mitigating ARI risk among the most vulnerable populations.

Geographic disparities in ARI prevalence were also observed (Table 3), with the highest rates in Province 6 (now Karnali Province) (3.37%) and Province 1 (now Koshi Province) (3.31%). Although these differences were not statistically significant in the multivariate model, they highlight important structural inequities. The NDHS 2022 findings confirm that Karnali Province still experiences the highest ARI burden with 4% prevalence.^19^ This persistent geographic disparity suggests structural inequities in healthcare access, environmental risk factors, and socioeconomic conditions. Provinces with higher rates of poverty, greater reliance on biomass fuels, and limited healthcare infrastructure are likely to experience higher ARI prevalence. The relationship between poverty, biomass fuel use, and ARI is well-established, though more research with improved exposure assessment is needed to validate findings and inform interventions. ^17 21^

Contrary to expectations, maternal education was not significantly associated with ARI prevalence (Table 3). Maternal education often influences child health outcomes through improved caregiving and health-seeking behaviors.^22 23^ This effect may be attenuated in Nepal due to broader systemic barriers, such as limited healthcare access and environmental risk factors. Community-based interventions, including health education programs can improve parental knowledge and awareness of ARI prevention and management, are crucial.^24^ Similarly, no significant differences in ARI prevalence were observed between urban and rural settings. This lack of association may be explained by urban areas facing unique challenges, including overcrowding, inadequate ventilation, and exposure to ambient air pollution, which offset the benefits of urbanization.

This study is strengthened by its use of a nationally representative dataset and rigorous statistical methods, including adjustments for the survey’s complex sampling design. However, certain limitations must be acknowledged. First, ARI status was based on caregiver-reported symptoms, which may introduce recall bias or misclassification. Second, the cross-sectional design precludes causal inferences. Third, the absence of data on critical risk factors, such as direct measures of household air pollution exposure and nutritional status, limited the scope of the analysis.

Despite these limitations, the findings underscore the critical role of socioeconomic and geographic disparities in shaping ARI risk among children in Nepal. Targeted public health interventions that address poverty, promote clean energy adoption, and enhance access to affordable healthcare are essential to mitigating the ARI burden. Improving access to quality healthcare services, including timely diagnosis, appropriate treatment, and effective case management, is essential.^2 25^ This involves strengthening health systems, training healthcare providers, and promoting appropriate healthcare-seeking behavior among parents.

Future research should focus on longitudinal studies to establish causal pathways and incorporate additional contextual factors, such as environmental exposures, healthcare quality, and cultural practices. Policy makers should consider these variables when taking steps to improve the health of children under 5 years of age who are affected by ARI. It is also important to keep regional variations in mind.

## Conclusion

Despite progress in reducing ARI prevalence from 2016 to 2022, disparities in geographic and socioeconomic determinants persist. Children from the poorest households faced a higher risk of ARI, emphasizing the need for poverty alleviation, clean energy alternatives, and improved living conditions. While caregivers demonstrated high healthcare-seeking behavior, the reliance on private facilities suggests inequities in access to affordable, quality care. Strengthening the public healthcare system and implementing region-specific interventions are critical to reducing ARI prevalence. Further research exploring environmental, nutritional, and healthcare factors is essential to inform comprehensive, evidence-based policies.

## Data Availability

The Demographic and Health Survey (DHS) Program, USA, is the custodian of the 2016 NDHS data for Nepal, which is freely available upon reasonable request submission to the DHS.

https://dhsprogram.com/methodology/survey/survey-display-472.cfm

## Acknowledgement

We heartly acknowledge the Demographic and Health Survey (DHS) Program, USA, for approving access to the NDHS 2016 data for Nepal.

## Authors’ contributions

Anish Dhodari developed the research question and study protocol, provided guidance for data analysis and conducted the data analysis.Anisha Dhakal, Joshna Pokhrel and Pragya Bhattarai drafted the initial manuscript. Anish Dhodari further revised and finalized the manuscript. All authors read and approved the final version.

## Declarations

### Ethics Approval and Consent to Participate

Not applicable.

### Consent for Publication

Consent was obtained from the DHS Program when accessing the data.

